# Monitoring compliance to topical therapy in children and young people with uveitis

**DOI:** 10.1101/2022.07.13.22277585

**Authors:** EKY Green, O McGrath, L Steeples, J Ashworth

**Affiliations:** Manchester Royal Eye Hospital, Oxford Road, Manchester, M13 9WL, UK; Roche Products UK Limited, 6 Falcon Way, Shire Park, Welwyn Garden City, AL7 1TW, UK; Manchester Royal Eye Hospital, Oxford Road, Manchester, M13 9WL, Division of Evolution & Genomic Sciences, University of Manchester, Manchester, UK

**Keywords:** Paediatric uveitis, children and young people with uveitis, adherence, compliance, eye drops, concordance

## Abstract

**Background:** Uveitis in children and young people is a rare but potentially debilitating condition. Steroid eye drops are the first step in treatment and poor compliance may result in vision-threatening complications. This study aimed to measure compliance with prescribed eye drops prospectively and investigate reasons for non-compliance in a child-specific manner.

**Methods:** 50 patients aged 0-18 years attending a tertiary paediatric uveitis clinic using steroid drops were recruited. Both the child or young person, and parent completed questionnaires about compliance. A subgroup had bottles of Prednisolone 1% drops weighed and dispensed at the first appointment and reweighed at follow-up. The reduction in weight was compared with expected weight change over the interval.

**Results:** The study was completed by 31 patients. Drop errors for all eye drops were reported more than “once a week” by 13 (33.3%) children and young people, and 3 (9.7%) parents. A large proportion of parents could not recall the frequency of drops prescribed to their child (13, 40.6%). Insufficient bottle weight reduction was found in 9 (75%), although this was not associated with self-reported drop errors. Within the eye drop weighing subgroup three (25%) used <50% the expected weight of drops.

**Conclusions:** Eye drop compliance was demonstrated to be poor by all the study outcomes in children and young people with uveitis. Self-reported compliance is unreliable in this population, as subjective responses were not supported by objective bottle weight change. Worryingly, some patients miss more than 50% of drops and may suffer worse disease control.

**Clinical implications:**

- **What is already known on this topic**
  - Poor drop compliance among children and young people with uveitis has been suggested by a previous retrospective study looking at self-reported compliance.
- **What this study adds**
  - All subjective and objective metrics in this prospective study suggest steroid eye drop compliance is often poor among children with uveitis.
  - Parental administration of steroid eye drops and shorter disease duration appear to be associated with improved compliance.
- **How this study might affect research, practice or policy**
  - Self-reported compliance is highly unreliable and should be treated as such by practising clinicians and any future studies.
  - Among the patients with drop errors a small but clinically significant number have very poor compliance and miss drops daily. These patients are at high risk for sub-optimal disease control or persistent disease activity and this requires further investigation.

**Synopsis/Precis:** This prospective study of drop compliance in children and young people with non-infectious uveitis suggests drop compliance is poor. Some individuals use drops less than half of the prescribed frequency. Self-reported compliance was highly unreliable.

## INTRODUCTION

Uveitis in children and young people remains a potentially sight-threatening condition, despite recent advancements in treatment,[1]. Topical steroids are first line therapy and a fundamental part of treatment of non-infectious uveitis. Children may also require intraocular pressure lowering drops for secondary raised intraocular pressure. Poor compliance with prescribed treatment may result in loss of vision. This can occur due to uncontrolled ocular inflammation and complications such as posterior synechiae, band keratopathy, cataract and secondary glaucoma.

Whilst it is acknowledged that poor compliance may lead to poor clinical outcomes, there is limited research into what barriers exist among children and young people with uveitis. Children and young people with uveitis face unique challenges; many cannot self-administer medication, the eye drop frequency may change week to week and there may be difficulties administering drops at school. The burden of disease can be substantial for the patient and their family due to treatment and the need for frequent follow-up. Treatment can include frequent eye drops and sometimes systemic medication.

Only one study has previously retrospectively assessed compliance in children and young people with uveitis,[2]. They found an association between frequency of eye drops, and parental drop administration and compliance. However, these findings may have been influenced by recall or assessor bias because compliance was assessed retrospectively based on notes taken by the same clinician that assessed disease activity.

The aim of the study is to quantify the range of adherence to eye drop medication prospectively, and investigate some of the reasons for non-compliance in a child and adolescent-specific manner.

## METHODS

Fifty patients with non-infectious uveitis were recruited from a Paediatric Uveitis clinic at the Manchester Royal Eye Hospital. Patient demographics were recorded including age, date of diagnosis and initiation of treatment, date of last change in treatment and systemic medication. At follow-up questionnaires were administered to children and people with parental responsibility (PPR) separately. The responses were then compared. The questionnaire was adapted from a questionnaire appropriate for children and young people designed for assessing compliance in paediatric glaucoma,[3]. The questionnaire asked the respondent to report the frequency of drops prescribed, and the frequency of errors for all prescribed drops since the time of recruitment. It also asked questions about difficulties encountered administering any drops.

Data was collected prospectively by a researcher different to the doctor assessing the patient’s clinical status. The questionnaires were administered to participants prior to their clinical assessment at the follow up appointment in order to minimise the risk of influencing participants’ responses.

A subgroup of patients had drop adherence assessed by change in Prednisolone 1% eye drop bottle weight. Patients had new bottles of eye drops weighed at the first appointment and again at their follow-up appointment. The drop weight change over the interval was compared to the expected bottle weight change if the drops had been administered correctly.

Ethics approval for this study was obtained from Health Research Authority, North East-Newcastle and North Tyneside 2 Research Ethics Committee according to the Declaration of Helsinki (REC reference 19/NE/0367). Participants gave informed consent to participate in the study before taking part. The study protocol was published on ClinicalTrials.gov (Identifier NCT04148365).

## RESULTS

A total of 50 patients were enrolled in the study, and each with a PPR; 31 children and young people and their respective PPR completed both questionnaires, 11 completed only one questionnaire (either a child or young person questionnaire, or a PPR questionnaire). Figure 1 contains the consort statement.

**Figure 1.**
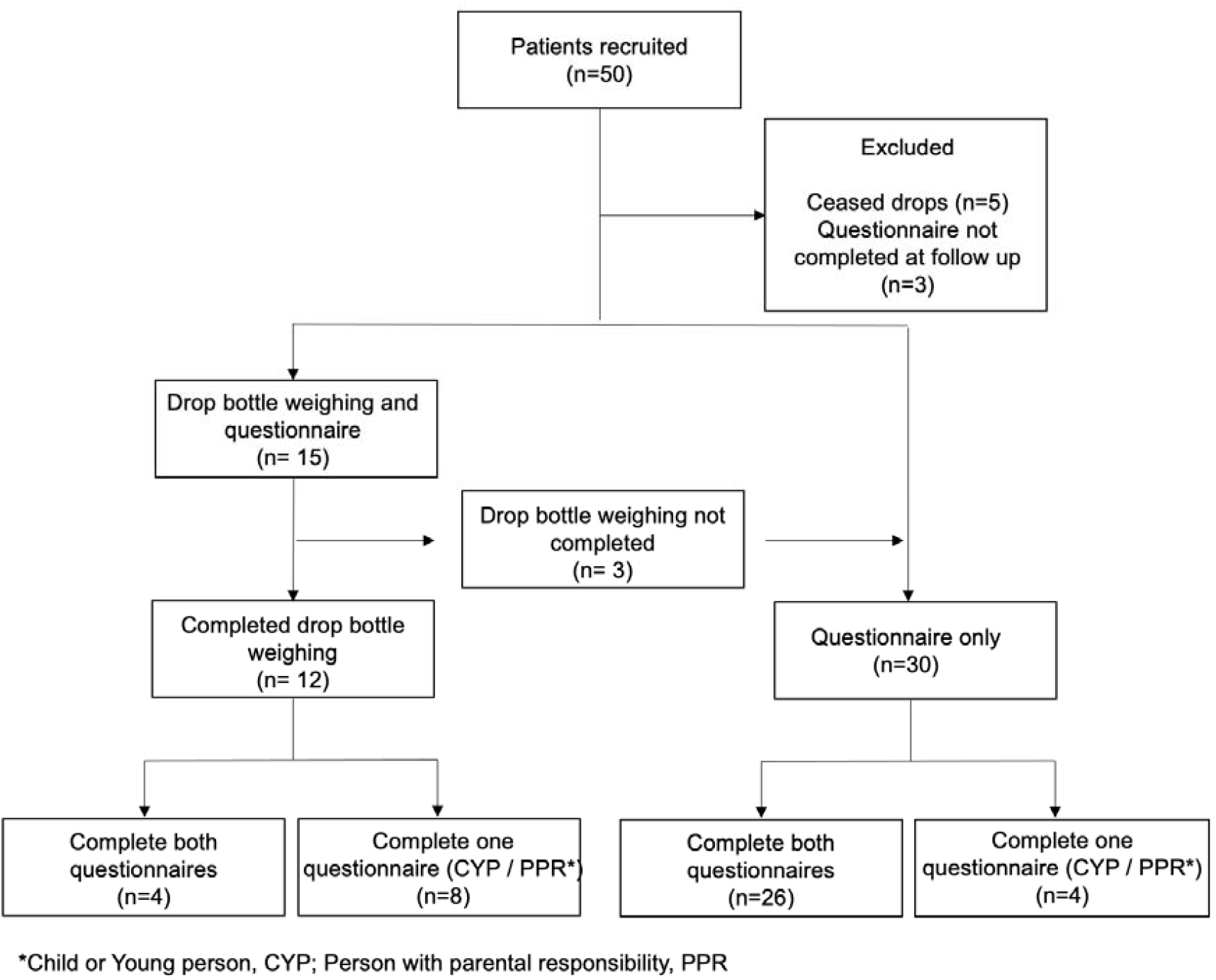
Consort statement.

The mean age of the child or young person was 12 years old (range 3-18 years), and 59.5% were female. The most common underlying diagnosis was Chronic idiopathic uveitis (n=28, 66.7%) (Table 1). Other diagnoses included Juvenile idiopathic arthritis (JIA) related uveitis (n=10, 23.8%), chronic uveitis secondary to trauma or surgery (n=2, 4.8%), idiopathic keratouveitis (n=1, 2.4) and Tubulointerstitial nephritis and uveitis (TINU) (n=1, 2.4). The ethnic origin varied widely but the majority recorded their background as white British (n=21, 51.2%), or Pakistani (n=5, 12.2%) (Table 1).

**Table 1.**
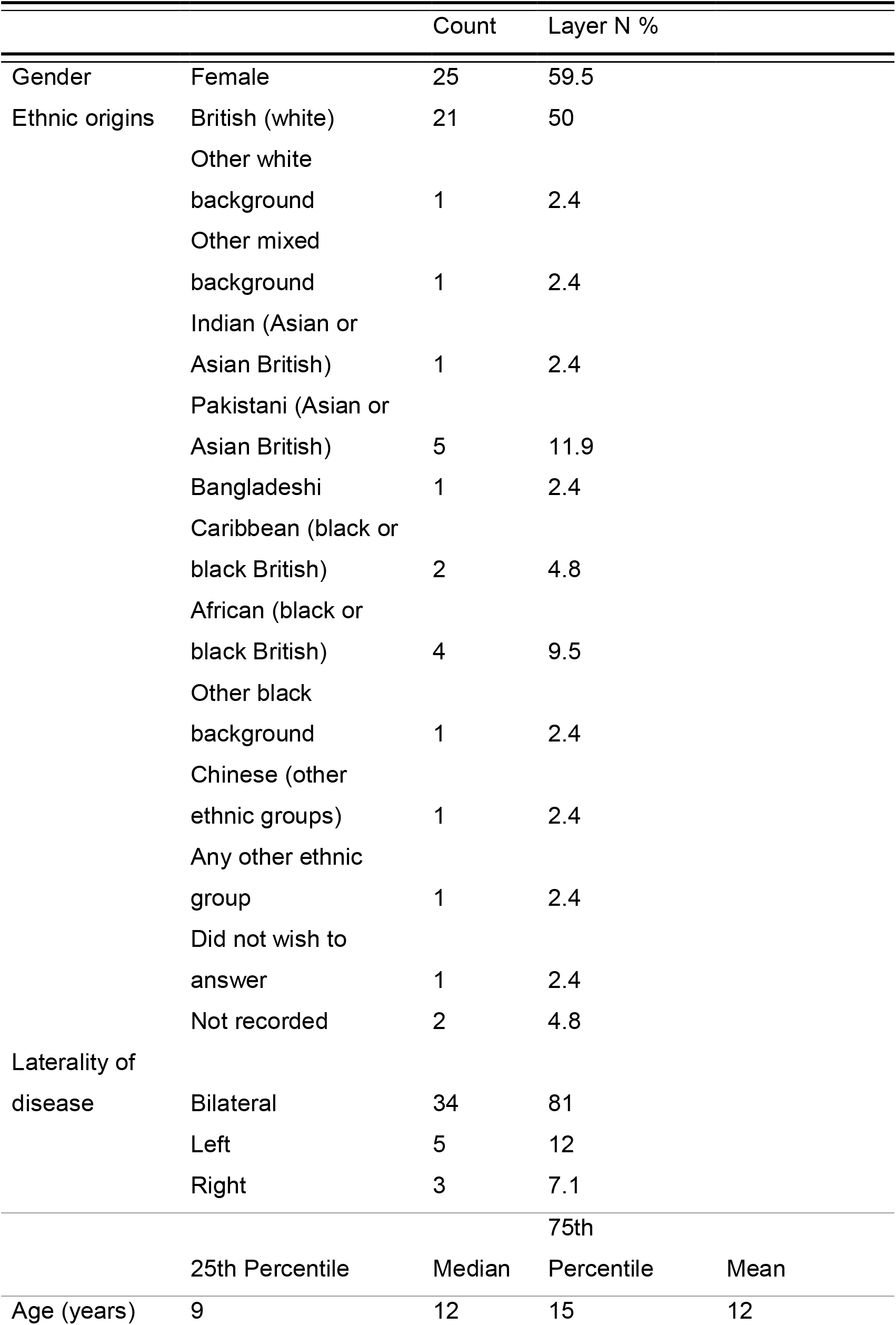
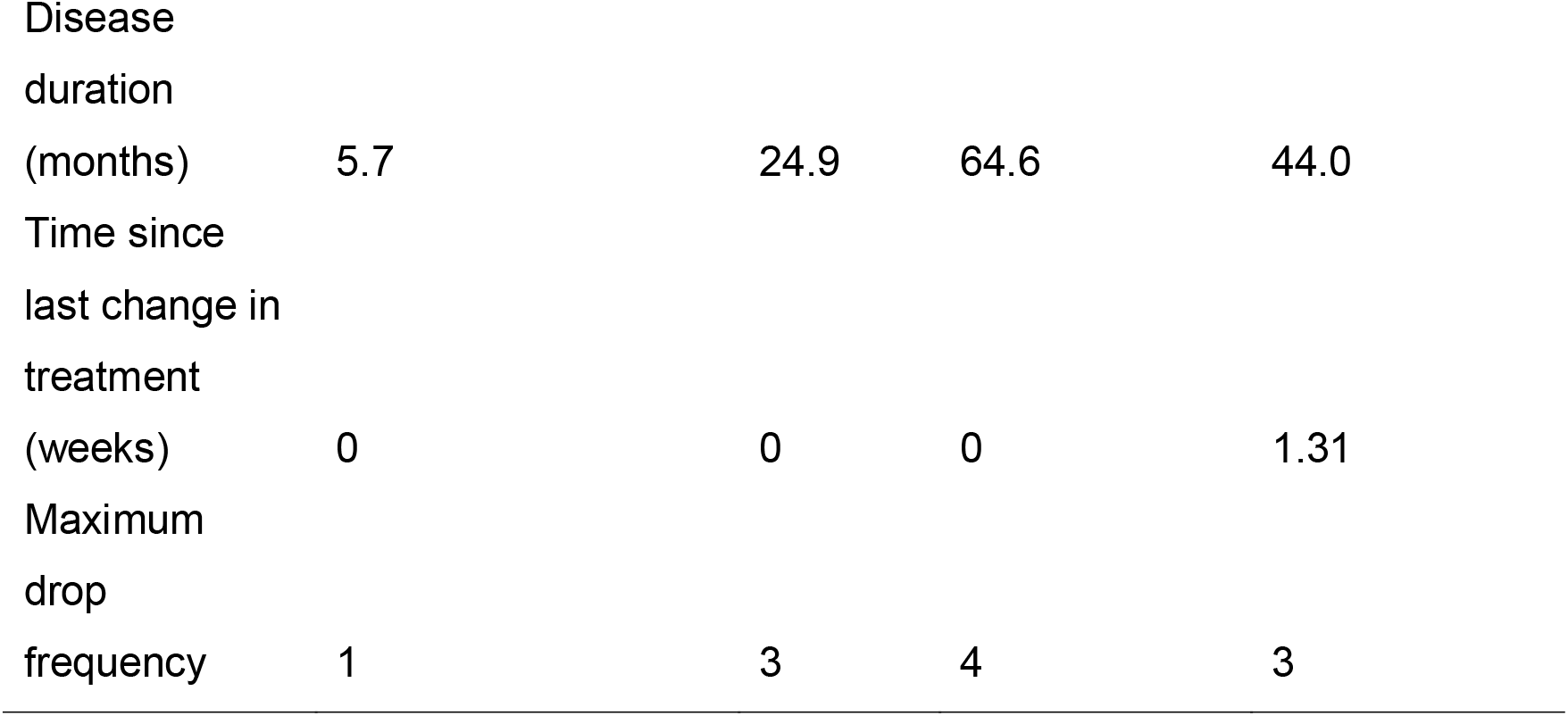

The mean time since diagnosis was 188 weeks (range 3 – 690 weeks). All patients had treatment initiated at the time of diagnosis and therefore no separate analysis between the duration of disease or duration of treatment can be performed. The median time since most recent change in medication was 0 weeks, although the mean time was 1.3 weeks (range 0-23 weeks). The mean maximum daily frequency of steroid eye drops was 3 (range 0.5-12). The mean total number of drops each person was prescribed was 1.73 (range 0.5-5). The drops prescribed to children and young people in this study included Prednisolone 1%, Cyclopentolate, Timolol 0.25% or 0.1%, Dexamethasone 0.1%, Latanoprost, Sodium hyaluronate 0.2%, Dorzolamide and Timolol combination drop and Iopidine 0.5%.

### Drop errors

Drop errors for all prescribed topical medication occurring “a few times a week” or more was reported by 13 (33.3%) children and young people, and 3 (9.7%%) PPR (Figure 2). When asked how often they were supposed to use the drops a different answer was given from the documented prescription by 12 (30.8%) children and young people, and 13 (40.6%) PPR.

**Figure 2.**
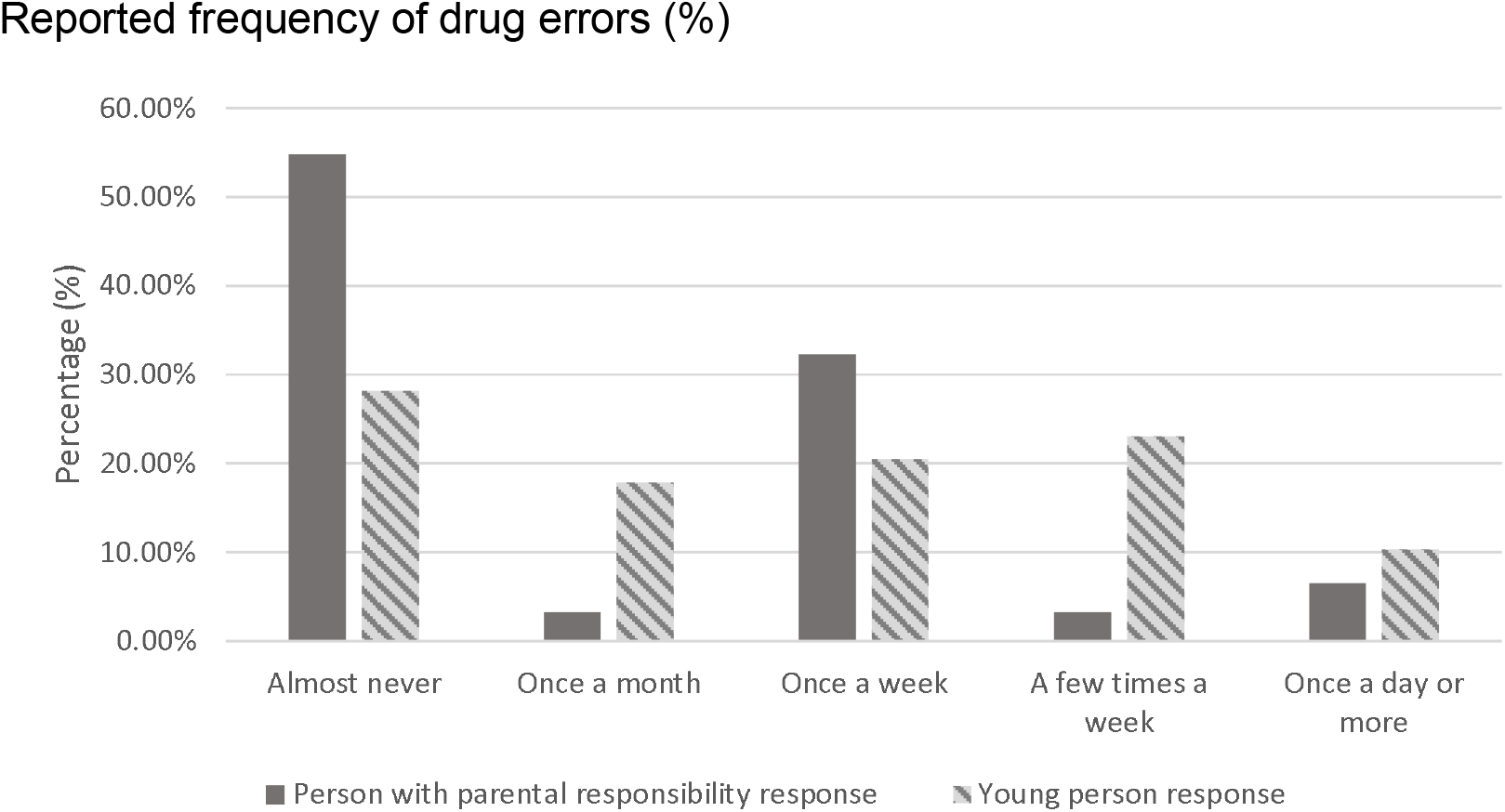
Reported frequency of drug errors (%)

### Factors associated with drop errors

No association was found between children and young people who reported missing drops “a few times a week or more” out of total prescribed drops and those who incorrectly recalled the prescribed drop frequency. All three adults that reported missing drops a few times a week or more also reported using drops differently from the prescription. However, there was an insufficient number of patients in each group to perform a test for significance.

In young people it was found that missing drops infrequently, “once a week or less,” was associated with also reporting a parent administering drops, and this was statistically significant (Fishers exact *p*=0.04). However, no association was found in parents responding to the same questions.

Duration of disease was lower in young people reporting missing drops “once a week” or less (Wilcoxon *p*= 0.001) (Figure 3).

**Figure 3.**
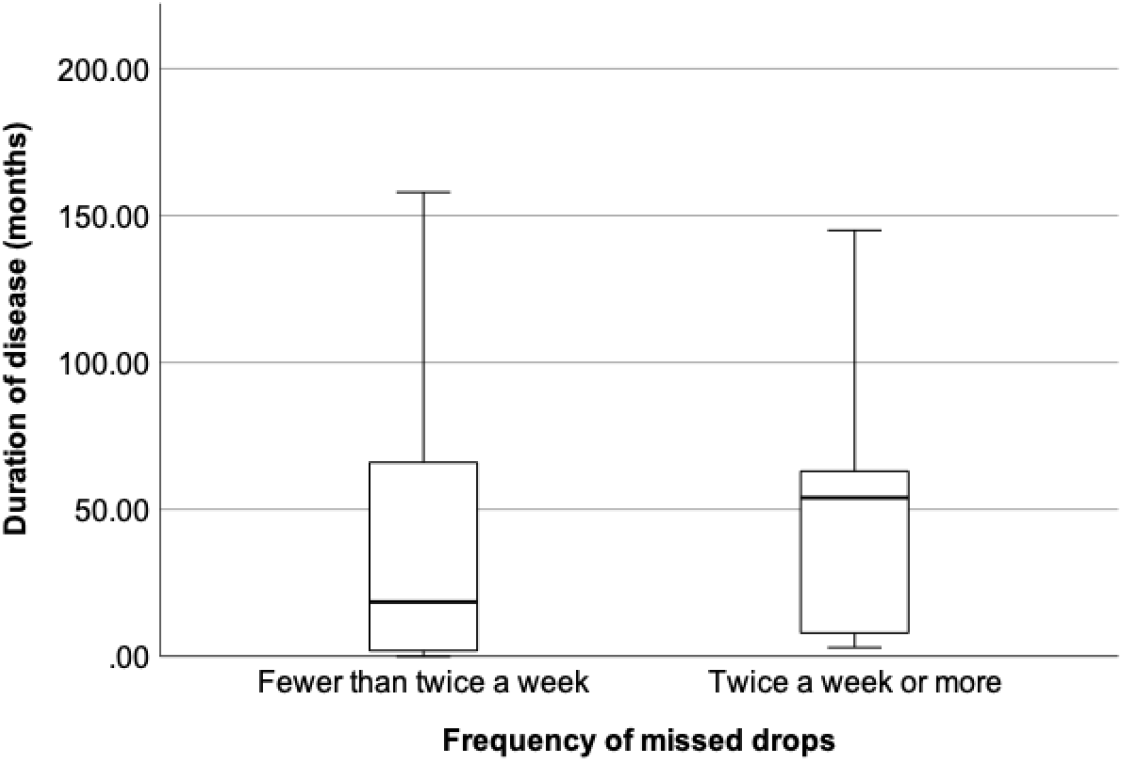

No association was found between the self-reported frequency of missing drops “a few times a week or more” and any other factor, including age under 11 years, gender, ethnicity, associated systemic disease, reported understanding of the disease and how important the respondent believed the drops were.

### Drop weight change

Prednisolone 1% eye drop bottles decreased in weight as expected or greater for 3 patients (25%). However, for the majority, the bottle weight reduced less than expected, suggesting that patients were missing doses (n = 9, 75%) (Figure 4).

**Figure 4.**
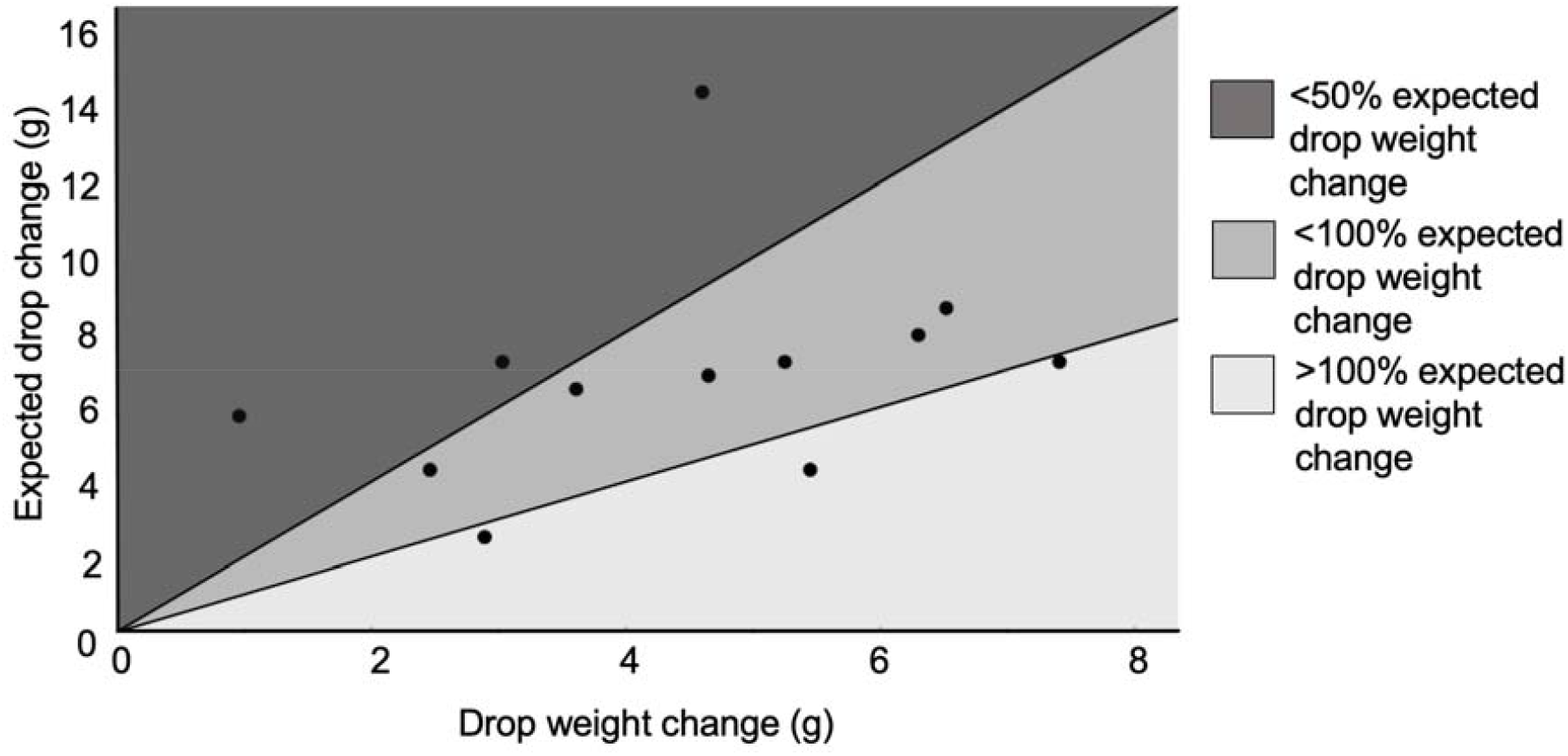
Comparison of recorded drop weight change with expected prednisolone 1% drop bottle weight change (g)

Among patients who appear to be under-using their Prednisolone 1% drops, three patients had a particularly low weight change that was less than half the expected weight change. These three patients were all 11 years or older and used drops 2-3 times daily. Two reportedly self-administered drops and one had a parent administer drops. All 3 were on systemic medication. When asked about how frequently they missed their drops one young person reported “once a month”, and two reported missing drops a “few times a week”. Only one parent completed the questionnaire, and they reported missing drops “once a week”. Of these patients, 2 children and young people reported prescribed drop frequency correctly, 1 incorrectly and the only parent responded correctly. Disease duration ranged from 11-276 weeks and current treatment duration ranged from 0-1 week. Disease duration and the entire duration of treatment were the same for all patients.

This differed from the three patients whose Prednisolone 1% drop bottle weight reduction was greater than expected, suggesting good compliance. These three individuals and their parents all reported the drop frequency correctly and reported that their parents administered the drops even though the age of the participant ranged from 9-14 years. There was no difference in ethnicity between groups. The current treatment regime duration was 0 weeks for all three, and disease duration ranged from 27-291 weeks.

### COVID-19

Data collection was interrupted and later resumed due to changes to clinic structure and restrictions on research during the pandemic period. Analysis to see if responses varied before and after the gap showed no difference. We performed an analysis to see if drug errors were reported more or less frequently when data was collected during COVID related school closures compared to data collected after schools reopened. No difference was found in the proportion of responses between the two periods.

## LIMITATIONS

Investigating medication compliance is problematic. Participants may inaccurately report their medication use either intentionally or unintentionally. They may also alter their compliance when under scrutiny.

To investigate the reliability of our measure of drop-error frequency, a comparison was performed between parental and child and young person responses. In 5 (16.7%) instances, the responses contradict each other, although most support each other.

We also compared self-reported drop errors with Prednisolone 1% bottle weight reduction. No association was found between bottle weight reduction and self-reported frequency of all missed drops. We investigated whether this was due to a misunderstanding of prescribed drop frequency and found no association between incorrect recollection and bottle weight change. This suggests that the problem is that self-reporting is unreliable.

Change in Prednisolone 1% bottle weight provides limited information about drop use. For example, patients may waste drops without using them, so even when bottle weight loss is in the expected range, it may be due to waste of drops rather than correct usage.

No indirect compliance monitors were included, such as uveitis flare-ups or recovery. This is because we observed a small number of patients over a variable window of time. Future studies would benefit from linking compliance with clinical outcomes in uveitis in children and young people.

## DISCUSSION

All the metrics used to evaluate drop compliance in this study in children and young people with uveitis suggest that large proportions of the cohort had poor compliance. This includes 33.3% of young people and 9.7% of responsible adults who reported total drop errors out of all prescribed drops more than once a week, 40.6% of adults who were unable to recall the correct prescribed frequency, and 75% of patients who had an insufficient weight reduction in their Prednisolone 1% eye drop bottles. However, we were unable to reliably quantify the size of the problem because the various metrics did not support each other.

Previous studies of paediatric glaucoma drop treatment found adult reports of drop use in children and young people to be reliable if the adult is administering the drops,[4]. This differs from our findings that children and young people were better at recalling drop frequency, and reported drop errors more frequently. The study by Moore et al. found that parents rarely reported non-adherence, in keeping with our findings,[4]. Wang et al. found that parents overestimated compliance with atropine for myopia,[5]. Self-reported adherence may be the only tool available in the average clinic. However, for research, more accuracy is required to make solid conclusions. Prednisolone 1% bottle weights revealed that among the patients that have poor compliance, there is a subgroup that has very poor drop adherence. Less than 50% adherence to medication would be expected to impact clinical outcomes. This is an area that would benefit from future research.

We identified two additional possible associations that might benefit from further investigation. Disease duration was negatively associated with self-reported drop errors for all drops used. Disease duration and the entire duration of treatment were the same for all patients and so the effect of one cannot be distinguished from the other. This suggests that people may lose motivation when the condition or treatment becomes longstanding. We are not aware of any studies reporting similar findings.

In contrast to a previous retrospective study into compliance among children and adolescents with uveitis we did not find any association between compliance and the daily frequency of eye drops,[2].

Drop administration by parents may be positively associated with compliance. The three patients who showed good compliance based on Prednisolone 1% bottle weight change had drops administered by parents even though they were all over 11 years of age, an age when the child or young person could theoretically self-administer drops. Previous studies have also shown compliance with paediatric glaucoma drops and uveitis drops was positively associated with parental administration,[2,3].

The absence of a single consistent factor affecting compliance such as age, drop frequency or reducing regime suggests that this is a complex subject. Drop frequency may be problematic for some patients but not all, and difficulties may be specific to particular children and young people and their environment. This study’s sample size may be too small to detect these trends.

## CONCLUSION

All metrics suggest drop compliance in children and young people with uveitis is often poor. This is clinically significant as poor drop compliance results in poorer disease control and an increased risk of subsequent complications due to uveitis. The majority of patients’ Prednisolone 1% bottle weights did not reduce as expected suggesting adherence is imperfect. Among this group, a subgroup has very poor compliance, and these patients may have persistent disease activity or sub-optimal control. Some possible associations with good compliance included parental help and a shorter disease duration. More research is needed to confirm these findings and identify what factors lead to poor compliance. Future research should not be based exclusively on self-reported adherence, which this study found had limited validity.

## Data Availability

All data produced in the present study are available upon reasonable request to the authors

## Abbreviations

PPR: Person with parental responsibility
JIA: Juvenile idiopathic arthritis
TINU: Tubulointerstitial nephritis and uveitis
CYP: Child or young person

## ACKNOWLEGEMENTS

We are grateful for funding from the British and Irish Paediatric Ophthalmology and Strabismus Association (BIPOSA). Thank you also to Rachel Harwood for statistical support.

